# Utility of cerebrovascular imaging biomarkers to detect cerebral amyloidosis

**DOI:** 10.1101/2024.05.28.24308056

**Authors:** Matthew D. Howe, Megan R. Caruso, Masood Manoochehri, Zachary J. Kunicki, Sheina Emrani, James L. Rudolph, Edward D. Huey, Stephen P. Salloway, Hwamee Oh, the Alzheimer’s Disease Neuroimaging Initiative

**Affiliations:** Butler Hospital Memory & Aging Program; 345 Blackstone Boulevard, Providence, RI, 02906, USA; Department of Psychiatry and Human Behavior, Brown University; 345 Blackstone Boulevard, Providence, RI, 02906, USA; University of Pennsylvania Perelman School of Medicine; 3400 Civic Center Blvd, Philadelphia, PA, 19014, USA; Center of Innovation in Long-Term Services and Supports, Providence VA Medical Center; 830 Chalkstone Ave, Providence, RI, 02908, USA; Department of Medicine, The Warren Alpert Medical School of Brown University; 222 Richmond Street, Providence, Rhode Island, 02903, USA

**Keywords:** ADNI, Alzheimer disease, amyloid-β, cerebrovascular disease, small vessel disease, magnetic resonance imaging, positron emission tomography

## Abstract

**INTRODUCTION:** The relationship between cerebrovascular disease (CVD) and amyloid-β (Aβ) in Alzheimer disease (AD) is understudied. We hypothesized that magnetic resonance imaging (MRI)-based CVD biomarkers, including cerebral microbleeds (CMBs), ischemic infarction, and white matter hyperintensities (WMH), would correlate with Aβ positivity on positron emission tomography (Aβ-PET).

**METHODS:** We cross-sectionally analyzed data from the Alzheimer’s Disease Neuroimaging Initiative (ADNI, N=1,352). Logistic regression was used to calculate odds ratios (ORs), with Aβ-PET positivity as the standard-of-truth.

**RESULTS:** Following adjustment, WMH (OR=1.25) and superficial CMBs (OR=1.45) remained positively associated with Aβ-PET positivity (p<.001). Deep CMBs and infarcts exhibited a varied relationship with Aβ-PET in cognitive subgroups. The combined diagnostic model, which included CVD biomarkers and other accessible measures, significantly predicted Aβ-PET (pseudo-R^2^=.41).

**DISCUSSION:** The study highlights the translational value of CVD biomarkers in diagnosing AD, and underscores the need for more research on their inclusion in diagnostic criteria. **ClinicalTrials.gov:** ADNI-2 (NCT01231971), ADNI-3 (NCT02854033)

## 1. BACKGROUND

Alzheimer disease (AD), the leading cause of dementia worldwide, is biologically defined by the presence of amyloid-β (Aβ) plaques and tau neurofibrillary tangles [1,2]. While understudied, neuropathological and neuroimaging studies indicate that cerebrovascular disease (CVD) is a common finding in AD patients, with AD and CVD co-pathology accounting for roughly 20-30% of cases in some cohorts [3–5]. The rising incidence of AD, partly attributed to increased life expectancy, is also driven by modifiable CVD risk factors, including hypertension, diabetes mellitus, and hyperlipidemia [6–8]. These modifiable risk factors have been shown to interact with age, sex and apolipoprotein-E ε4 (APOE-ε4) genotype, potentially influencing susceptibility to AD via actions on glial cells, changes in blood-brain barrier structure/function, or perivascular clearance of Aβ peptides [7,9–11]. Much interest has been paid to using structural neuroimaging to identify small vessel disease and vascular dementia *in vivo*, with the dominant scales focused on detection of vascular Aβ deposition rather than total Aβ burden (i.e. parenchymal and vascular plaques) that is considered to be the hallmark of AD [2,7,12–14]. Therefore, more work is needed to characterize the potential utility of CVD biomarkers in identifying Aβ pathology more broadly [5,7].

Seeking to identify a cerebrovascular “signature” of Aβ pathology in AD, the current study explores the utility of existing magnetic resonance imaging (MRI)-based CVD biomarkers. We investigated the degree to which CVD neuroimaging biomarkers—including cerebral microbleeds (CMBs), asymptomatic lacunar infarction, and white matter hyperintensities (WMH)—correlate with Aβ positivity as determined by positron emission tomography (Aβ-PET). We selected these CVD biomarkers due to: (1) high-quality neuropathological evidence linking them to relevant age-related cerebral small vessel diseases and/or neuropathology of AD (i.e. Thal, Braak or CERAD staging) [13], (2) the utility of incorporating these biomarkers into existing algorithms for small vessel disease detection, such as the Boston Neuroimaging Criteria 2.0 or the Cerebral Small Vessel Disease Score [4,12,14], as well as (3) relative ease of assessment by physicians in routine practice with clinical MRI protocols [15]. This approach is further supported by the Amyloid/Tau/Neurodegeneration framework which allows for biomarker confirmation of Aβ positivity (i.e. Aβ-PET) to biologically define AD [2].

Based on the existing literature, our hypothesis is twofold: first, that individual CVD biomarkers will differentially predict Aβ-PET positivity, and second, that their inclusion, alongside other accessible clinical, genetic and neuroimaging measures, will improve multimodal prediction of Aβ-PET positivity [16–21]. To test these hypotheses, we used participant data from the Alzheimer’s Disease Neuroimaging Initiative (ADNI), a well-described observational cohort study of older adults with AD [22,23]. Taking advantage of this cohort enriched for AD, we primarily sought to characterize the associations between commonly identified MRI-visible CVD measures and Aβ-PET and examine their diagnostic performance within an accessible algorithm which includes demographic (age, sex), genetic (APOE-ε4 status) and cognitive (Montreal Cognitive Assessment [MoCA]) measures that were available in the dataset [22,23]. To enhance the translational value of our results, we additionally stratified patients by cognitive stage for all analyses—informing their potential usefulness for cognitively unimpaired (CU), mild cognitive impairment (MCI), and dementia.

## 2. METHODS

### 2.1 Participants

The ADNI was launched in 2003 as a public-private partnership, led by Principal Investigator Michael W. Weiner, MD. This study utilizes data obtained from ADNI-2 (NCT01231971) and ADNI-3 (NCT02854033), two recent phases of a landmark study that has collected data at over 50 sites in North America and has been used to develop clinical, imaging, genetic, and biochemical biomarkers for the early detection and tracking of AD over the past 20 years (adni.loni.usc.edu) [22,23]. The primary goal of ADNI has been to test whether serial MRI, PET, other biological markers, and clinical and neuropsychological assessment can be combined to measure the progression of MCI and early AD.

Participants in ADNI are aged 55-90, and either English or Spanish-speaking. Exclusions include inability to tolerate a blood draw, contraindication to neuroimaging, evidence of infarction or other focal brain lesion (those with multiple lacunes or lacunes in a critical memory structure are excluded), any significant neurologic disease, major recent psychiatric disorders, or significant systemic illness or medical conditions. Baseline diagnoses were determined using a combination of cut-off scores from several instruments including the Mini-Mental Status Examination (MMSE), Cognitive Change Index (CCI), Weschler Logical Memory Test, and Clinical Dementia Rating (CDR®) Staging Instrument. Based on these assessments, participants were assigned a diagnostic category based on expert consensus of Cognitively Normal (CN), Subjective Memory Complaint (SMC), Mild Cognitive Impairment (MCI), and AD Dementia. Written informed consent was obtained from all participants or authorized representatives prior to the start of study procedures, with local Institutional Review Boards and Research Ethics Boards providing oversight for the study. The authors certify that the study was performed in accordance with the ethical standards as laid down in the 1964 Declaration of Helsinki and its later amendments.

### 2.2 ADNI Dataset

For the current analysis, we downloaded demographic, neuroimaging and genetic data from the Laboratory of Neuro-Imaging (LONI) site where ADNI data is housed, including Aβ-PET, structural MRI and APOE genotype determined by the Illumina Human BeadChip panel [24]. Due to differences in imaging protocols and PET tracers in ADNI-1, we limited our analysis to participants who screened for ADNI-2 and ADNI-3 (N = 1,430). After review of screening and baseline visit data, 78 individuals with unknown Aβ-PET status (N = 76) or absent clinical diagnosis (N = 2) were excluded to yield our final sample (N = 1,352).

Dataframes were downloaded from LONI using the *adnimerge* R package *(version 0.1.1)*. We obtained up-to-date demographic, cognitive and apolipoprotein-E genotypes from the Alzheimer’s Disease Cooperate Study *(ADNIMERGE).* Additionally, we obtained imaging data including whole and regional brain volumes from University of California at San Francisco *(ADNIMERGE),* WMH and infarct data from the Center for Neuroscience at University of California at Davis *(UCD_WMH_05_02_22, MRI_INFARCTS_01_29_21),* region-specific CMB counts from the Mayo Clinic Aging and Dementia Imaging Research Laboratory *(MAYOADIRL_MRI_MCH_09_07_22),* as well as florbetapir and florbetaben standardized uptake value ratios (SUVR) summary data from the University of California at Berkley *(UCBERKELEYAV45_04_26_22, UCBERKELEYFBB_04_26_22)*. For a detailed summary of the standard imaging acquisition and analysis pipelines used in ADNI, see **Supplementary Methods**. We last accessed the ADNI database on 27 December 2023.

### 2.3 Determination of Aβ-positivity

Participants were classified as Aβ-positive or Aβ-negative based on Aβ-PET standardized uptake value ratios (SUVR) according to the standard analysis pipeline at the University of California at Berkley (see **Supplementary Methods**). Aβ-PET positivity was defined as SUVR > 1.11 and SUVR > 1.08 for florbetapir and florbetaben, respectively [25,26]. For quantitative comparisons, SUVR was transformed to a standardized centiloid scale as previously described by Royse et al. [27].

### 2.4 Quantification of medial temporal lobe and white matter volumes

Using T1 and T2-weighted MRI images, WMH and medial temporal lobe (MTL) volumes were quantified by the Center for Neuroscience at University of California at Davis and the University of California at San Francisco, respectively, according to the standard Freesurfer analysis pipeline (See **Supplementary Methods**). To control for inter-individual and disease stage-related variability in global atrophy, mean MTL and total WMH volumes were normalized to whole brain volumes, then log transformed.

### 2.5 Quantification and localization of CMBs

“Definite” or “probable” CMBs were identified and reported by the Mayo Clinic Aging and Dementia Imaging Research Laboratory by review of T2* gradient recall echo images according to the standard analysis pipeline (see **Supplementary Methods**). We divided CMBs into lobar (cortical and cerebellar) or deep (subcortical, periventricular white matter and brainstem regions) as previously described by Charidimou et al. [12]. We then classified participants by the presence or absence of superficial and deep CMBs, respectively.

### 2.6 Quantification of lacunar infarcts

Lacunar infarcts were identified by a physician specifically trained in interpretation of MRI, with infarct size, location and other imaging characteristics recorded according to the standard analysis pipeline at the Center for Neuroscience at University of California at Davis (see **Supplementary Methods**). For this analysis, we only considered lesions equal to or greater than 3mm in size as cerebral infarcts. We then classified participants based on the presence or absence of infarcts [28].

### 2.7 Covariates

We adjusted all models for demographics (age, sex), cognition (MoCA score) and APOE genotype (number of ε4 alleles).

### 2.8 Statistics

Data cleaning, analysis and plotting was conducted in R using the *mice, psfmi, pROC* and *ggplot2* packages. Missing data comprised no more than 5% of the overall sample and were handled by multiple imputation by chained equations (**Supplementary Table 1**). Descriptive statistics were calculated for each group and reported as mean (SD), median (IQR) and N (%) according to Rubin’s Rules [29]. Adjusted odds ratios (OR) were calculated by logistic regression with dichotomized Aβ-PET status as the outcome, reported with a 95% confidence interval (CI), with statistical significance determined by the Wald test. For evaluation of regression model goodness-of-fit and parsimony, we used prediction model pooling, selection, and performance evaluation across multiply imputed datasets to calculate the Nagelkerke Pseudo-R^2^ and Akaike Information Criterion (AIC). The Hosmer-Lemeshow test was used to test goodness-of-fit compared to the null model, with p > 0.05 denoting acceptable model fit to the observed data.

Receiver operator characteristic (ROC) analysis was performed to calculate area under the curve (AUC) as well as diagnostic sensitivity, specificity and accuracy at Youden’s optimal cut-off. Nested models were created using forward variable selection: Step 1: Base (Age, Sex, APOE4 genotype), Step 2: Base + MTLv, and Step 3: Base +MTLv + CVD (see **Supplementary Table** 2 for further details on model components). Nested models with forced forward selection were compared using the likelihood ratio test for multiply imputed data, reported as the d3 statistic with degrees of freedom (df) corresponding to difference in number of predictors between model steps. Statistical significance was defined by a threshold of p < .05.

## 3. RESULTS

### 3.1 Participant characteristics

To assess the relationships between CVD biomarkers and the outcome (Aβ-PET positivity), we analyzed cross-sectional screening visit data for ADNI-2 and ADNI-3 (N = 1,352), then subdivided participants based on cognitive stage at screening (**Table 1**). For the purposes of our analysis, participants categorized at their screening visit as cognitively normal or experiencing subjective memory complaints were classified as cognitively unimpaired (CU, N = 611), encompassing 198 (32.4%) Aβ-PET positive and 413 (67.6%) Aβ-PET negative participants. The MCI cohort (N = 531) contained 294 (55.4%) Aβ-PET positives and 237 (44.6%) Aβ-PET negative participants. Finally, the dementia cohort (DEM, N = 210) was predominantly Aβ-PET positive (N = 181 [86.2%]) with a smaller proportion Aβ-PET negative participants (N = 29 [13.8%]) (**Table 1, Supplementary Table 1**).

**Table 1:** Baseline demographic and clinical characteristics.

|  | Cognitively Unimpaired |  | Mild Cognitive Impairment |  | Dementia |  |
| --- | --- | --- | --- | --- | --- | --- |
| Characteristic* | Aβ- (N = 413) | Aβ+ (N = 198) | Aβ- (N = 237) | Aβ+ (N = 294) | Aβ- (N = 29) | Aβ+ (N = 181) |
| Age, years | 70 (6) | 73 (7) | 70 (8) | 73 (7) | 76 (8) | 74 (8) |
| Sex, male | 184 (45%) | 72 (36%) | 134 (57%) | 160 (54%) | 24 (83%) | 100 (55%) |
| Education, years | 16.78 (2.34) | 16.53 (2.48) | 16.35 (2.46) | 16.29 (2.61) | 16.62 (2.24) | 15.51 (2.61) |
| APOE-ε4, alleles |  |  |  |  |  |  |
| 0 | 313 (77%) | 97 (49%) | 180 (78%) | 91 (31%) | 23 (79%) | 47 (26%) |
| 1 | 87 (21%) | 89 (45%) | 46 (20%) | 151 (52%) | 6 (21%) | 88 (49%) |
| 2 | 6 (1.5%) | 11 (5.6%) | 5 (2.2%) | 51 (17%) | 0 (0%) | 43 (24%) |
| MoCA, score | 26.05 (2.58) | 25.65 (2.51) | 23.7 (3.0) | 22.5 (3.3) | 18.2 (4.0) | 16.9 (4.6) |
| MTLv, normalized (log) | -4.21 (0.10) | -4.22 (0.10) | -4.23 (0.10) | -4.28 (0.11) | -4.31 (0.13) | -4.38 (0.12) |
| WMHv, normalized (log) | -6.84 (1.33) | -6.24 (1.42) | -6.47 (1.40) | -6.03 (1.36) | -6.02 (1.01) | -5.69 (1.13) |
| CMBs, any location (1+) | 84 (21%) | 47 (24%) | 58 (25%) | 95 (32%) | 6 (21%) | 73 (41%) |
| CMBs, superficial (1+) | 61 (15%) | 42 (21%) | 45 (19%) | 82 (28%) | 5 (18%) | 66 (37%) |
| CMBs, deep (1+) | 27 (6.6%) | 10 (5.1%) | 12 (5.1%) | 27 (9.2%) | 1 (3.6%) | 18 (10%) |
| Infarct (1+) | 24 (6.8%) | 22 (12%) | 17 (8.7%) | 23 (9.1%) | 1 (4.3%) | 7 (4.5%) |
| Aβ-PET burden, cI<sup>†</sup> | 4 (9) | 55 (31) | 1 (10) | 73 (33) | -3 (14) | 88 (30) |

\*Mean (SD); n (%); Median (IQR); <sup>†</sup>cI = centiloids See **Supplementary Table 1** for missing values.

### 3.2 Development of the adjusted model for prediction of Aβ-PET positivity

We next sought to gauge the relationship between specific MRI markers of CVD neuropathology and Aβ-PET positivity in the overall cohort using a model that was adjusted by accessible measures of vascular risk (age, sex, APOE-ε4), cognitive impairment (MoCA) and AD-related atrophy (MTL volume [MTLv]) (**Table 2**). After adjustment for these covariates, we identified statistically significant effects of MRI-based CVD biomarkers. We found that increased WMHv (OR = 1.25 [95% CI: 1.20, 1.29], p < .001) and presence of one or more superficial CMBs (OR = 1.45 [95% CI: 1.31 – 1.61], p < .001) as being associated with Aβ-PET positivity in the overall cohort ( **Table 2**). We did not identify any statistically significant relationships between deep CMBs (OR = 0.88 [95% CI: 0.74 – 1.04], p = .12) or ischemic infarction (OR = 1.01 [95% CI: 0.89 – 1.15], p = 0.40) and Aβ-PET positivity in the overall cohort (**Table 2**). The adjusted model exhibited a pseudo-R^2^ = .41, consistent with a large effect size for goodness-of-fit.

**Table 2:** Prediction of Aβ-PET positivity in the overall cohort.

| Characteristic | OR<sup>*</sup> | 95% CI<sup>†</sup> | p-value |
| --- | --- | --- | --- |
| Age, years | 1.05 | 1.04, 1.06 | < .001 |
| Sex, male | 0.66 | 0.60, 0.71 | < .001 |
| APOE-ε4, alleles | 5.66 | 5.24, 6.11 | < .001 |
| MoCA, score | 0.89 | 0.88, 0.90 | < .001 |
| MTLv, normalized (log) | 0.12 | 0.08, 0.17 | < .001 |
| WMHv, normalized (log) | 1.25 | 1.20, 1.29 | < .001 |
| CMBs, superficial (1+) | 1.45 | 1.31, 1.61 | < .001 |
| CMBs, deep (1+) | 0.88 | 0.74, 1.04 | .12 |
| Infarction (1+) | 1.01 | 0.89, 1.15 | .90 |

\*OR = Odds Ratio, <sup>†</sup>CI = Confidence Interval. N = 1,352.

Based on these findings, we next sought to understand use this model to understand how the relationships between CVD biomarkers and Aβ-PET positivity may differ across different cognitive stages. Thus, we repeated this regression analysis in the CU, MCI and DEM cohorts, described in the following sections which are organized by CVD biomarker of interest.

### 3.3 Cerebral microbleeds

We hypothesized that the presence of superficial CMBs would be associated with Aβ-PET positivity across all cognitive stages. Examining each cohort separately, we identified a consistent and statistically significant relationship between superficial CMBs and Aβ-PET positivity, as observed in the CU (OR = 1.38 [95% CI: 1.18, 1.62], p < .001), MCI (OR = 1.37 [95% CI: 1.17, 1.61], p < .001), and DEM cohorts (OR = 2.17 [95% CI: 1.46, 3.29], p < .001; **Table 3**; **Figure 1, panel 1**).

**Figure 1:**
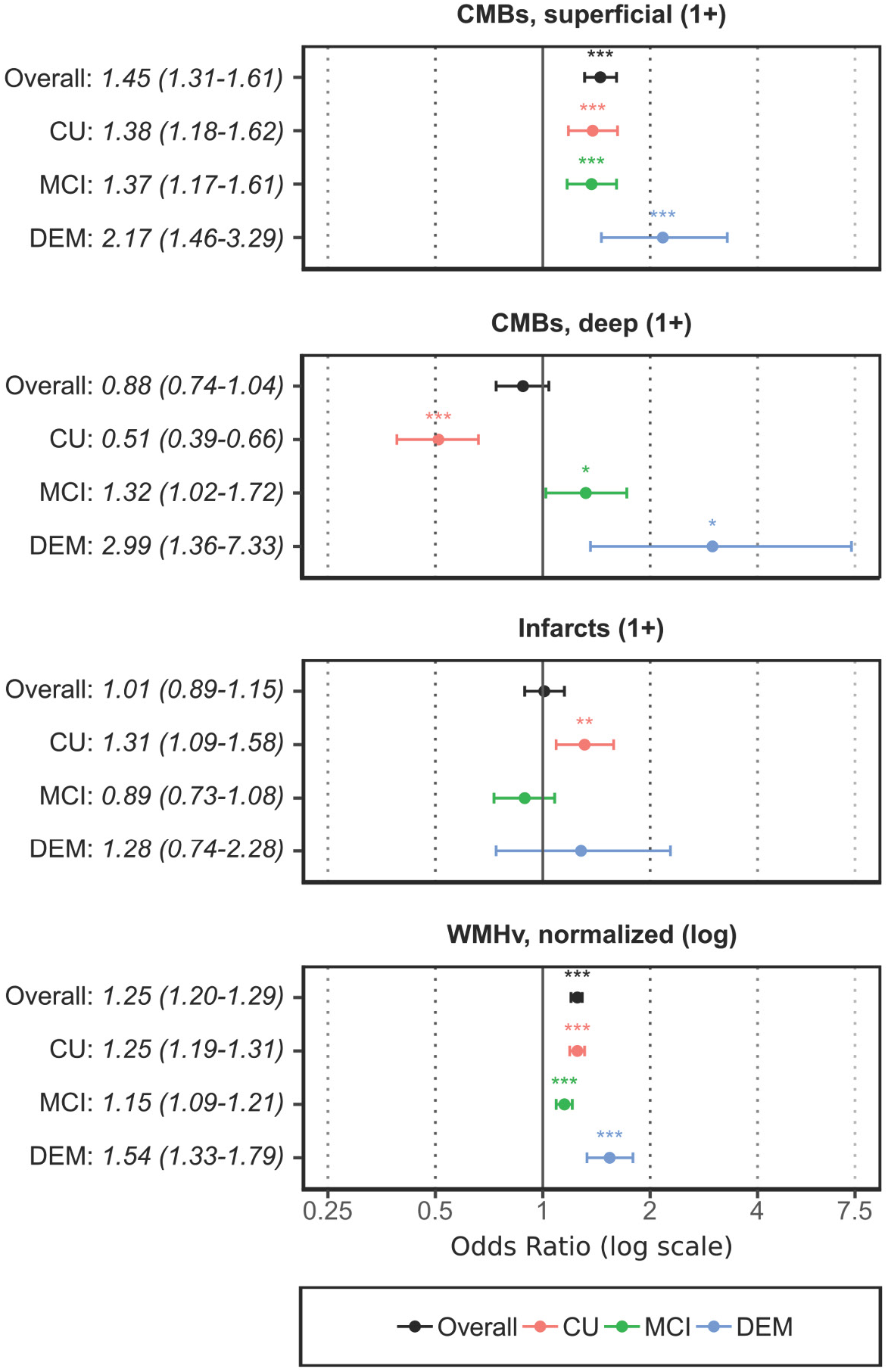
Differential prediction of Aβ-PET positivity by CVD-related MRI findings. Forest plots show the adjusted odds ratios (ORs) with 95% confidence intervals (CIs) for each cohort: Cognitively Unimpaired (CU, N = 611, red), Mild Cognitive Impairment (MCI, N = 531, green), and Dementia (DEM, N = 210, blue), compared to the overall effect (black). ORs were derived from logistic regression models adjusted for relevant covariates. Cerebral Microbleeds (CMBs) and White Matter Hyperintensity volume (WMHv) are shown to be significant predictors in varying degrees across cohorts. Ischemic infarcts were significant predictors in the CU cohort, only. Statistical significance determined by the Wald test is denoted as follows: *p < .05, **p < .01, ***p < .001.

**Table 3:** Adjusted cerebrovascular predictors of Aβ-PET positivity by cohort.

| Characteristic | OR* | 95% CI† | p-value |
| --- | --- | --- | --- |
| **CU Cohort (N = 611)** |  |  |  |
| CMBs, superficial (1+) | 1.38 | 1.18, 1.62 | < .001 |
| Infarction (1+) | 1.31 | 1.09, 1.58 | .005 |
| WMHv, normalized (log) | 1.25 | 1.19, 1.31 | < .001 |
| CMBs, deep (1+) | 0.51 | 0.39, 0.66 | < .001 |
| **MCI Cohort (N = 531)** |  |  |  |
| CMBs, superficial (1+) | 1.37 | 1.17, 1.61 | < .001 |
| CMBs, deep (1+) | 1.32 | 1.02, 1.72 | .037 |
| WMHv, normalized (log) | 1.15 | 1.09, 1.21 | < .001 |
| Infarction (1+) | 0.89 | 0.73, 1.08 | .20 |
| **Dementia Cohort (N = 210)** |  |  |  |
| CMBs, deep (1+) | 2.99 | 1.36, 7.33 | .011 |
| CMBs, superficial (1+) | 2.17 | 1.46, 3.29 | < .001 |
| WMHv, normalized (log) | 1.54 | 1.33, 1.79 | < .001 |
| Infarction (1+) | 1.28 | 0.74, 2.28 | .40 |

\*Adjusted Odds ratios (OR) were adjusted for age, sex, APOE genotype, cognition (MoCA) and MTL volume.
†CI = Confidence Interval.
See **Supplementary Table 3** for the full model with covariates.

Conversely, when examining deep CMBs, we hypothesized that they would not significantly predict Aβ-PET status for any cognitive stage. However, our analysis revealed differences in these relationships depending on cognitive stage: a negative association in the CU cohort (OR = 0.51 [95% CI: 0.39, 0.66], p < .001) contrasted with a positive association in the MCI (OR = 1.32 [95% CI: 1.02, 1.72], p = .037) and DEM cohorts (OR = 2.99 [95% CI: 1.36, 7.33], p = .011; **Table 3**; **Figure 1, panel 2**). These data indicate a potential complex relationship between deep CMBs and Aβ-PET that appears to be dependent on cognitive stage.

### 3.4 Ischemic infarction

We next sought to characterize how ischemic infarction might correlate with Aβ-PET positivity. Given the relatively low frequency of infarcts identified in our sample, we were unable to separately analyze infarct location or size-dependent effects. Regardless, analyzing each cognitive stage separately revealed a significant association of one or more infarcts in the CU cohort (OR = 1.31 [95% CI: 1.09, 1.58], p = .005), but not in the MCI (OR = 0.89 [95% CI: 0.73, 1.08], p = 0.200) or DEM cohorts (OR = 1.28 [95% CI: 0.74, 2.28], p = .400; **Table 3**; **Figure 1, panel 3**). There was significant variability, highlighting a heterogeneous relationship between ischemic lesions and Aβ-PET.

### 3.5 White matter hyperintensities

We next investigated the predictive value of WMHv (normalized to whole brain volume) for Aβ-PET positivity, postulating that the effects would be consistent across the disease course. We found that increased WMHv was a significant predictor of Aβ-PET positivity across all disease stages: CU (OR = 1.25 [95% CI: 1.19, 1.31], p < .001), MCI (OR = 1.15 [95% CI: 1.09, 1.21], p < .001), and DEM (OR = 1.54 [95% CI: 1.33, 1.79], p < .001; **Table 3**; **Figure 1, panel 4**). These results remained significant after adjustment for other CVD measures, and additional covariates, indicating a consistent and strong relationship between WMHv and Aβ-PET positivity across the cognitive spectrum, irrespective of MTL atrophy or whole brain volume.

### 3.6 Added benefit to assessing CVD biomarkers to predict Aβ-PET positivity

To evaluate the influence of cumulative CVD burden on the likelihood of Aβ-PET positivity, and to model how these biomarkers might be used in the context of a clinical work-up, we next employed nested logistic regression models in our analysis of the overall cohort. Step 1 (Base) included participant demographics (age, sex), genetic screening (APOE-ε4), and cognition (MoCA). Step 2 (Base + MTLv) included the same variables, with the addition of MTLv as a clinically relevant surrogate for AD-related atrophy. Finally, Step 3 (Base + MTLv + CVD) incorporated our CVD biomarkers of interest (Superficial and Deep CMBs, Infarcts, and WMHv). To compare the “added benefit” for each model step, we examined the change in AIC (Δ AIC) values between nested model steps, with negative values indicating improved parsimony in model prediction of Aβ-PET positivity (i.e. improved goodness-of-fit without substantially increasing model complexity). For a list of these variables for each model step in tabular format, see **Supplementary Table 2**.

In the overall cohort, our base model included age, sex, APOE4 genotype, and cognition (MoCA) as covariates (Step 1: pseudo-R^2^ = .38; **Table 4**). As expected, we found that assessing MTL volume (MTLv), a commonly used measure of atrophy, enhanced model fit (Step 2: pseudo-R^2^ = .39, Δ AIC = –11.43, d_3_[1] = 11.33, p = .001; **Table 4**). Importantly, further improvements in fit and parsimony were observed upon adding CVD biomarkers to the model, even when MTL atrophy had already been included (Step 3: pseudo-R^2^ = .41, Δ AIC = –15.78, d_3_[4] = 5.30, p < .001). These findings indicate that for the overall cohort, inclusion of CVD biomarkers produced additional benefits to model fit without increasing complexity (i.e., the model became more parsimonious).

**Table 4:** Prediction of Aβ-PET positivity with nested models in the overall cohort.

| Characteristic | Model 1: Base<br>(Pseudo-R<sup>2</sup> = .38) |  |  | Model 2: Base + MTLv<sup>†</sup><br>(Pseudo-R<sup>2</sup> = .39) |  |  | Model 3: Base + MTLv<sup>†</sup> + CVD<sup>2</sup><br>(Pseudo-R<sup>2</sup> = .41) |  |  |
| --- | --- | --- | --- | --- | --- | --- | --- | --- | --- |
|  | OR* | 95% CI<sup>†</sup> | p-value | OR* | 95% CI<sup>†</sup> | p-value | OR* | 95% CI<sup>†</sup> | p-value |
| Age, years | 1.08 | 1.07, 1.08 | < .001 | 1.07 | 1.06, 1.08 | < .001 | 1.05 | 1.04, 1.06 | < .001 |
| Sex, male | 0.67 | 0.62, 0.73 | < .001 | 0.66 | 0.60, 0.71 | < .001 | 0.66 | 0.60, 0.71 | < .001 |
| APOE-ε4, alleles | 5.70 | 5.29, 6.15 | < .001 | 5.63 | 5.22, 6.08 | < .001 | 5.66 | 5.24, 6.11 | < .001 |
| MoCA, score | 0.86 | 0.85, 0.87 | < .001 | 0.89 | 0.88, 0.90 | < .001 | 0.89 | 0.88, 0.90 | < .001 |
| MTLv, normalized (log) | - | - | - | 0.10 | 0.07, 0.15 | < .001 | 0.12 | 0.08, 0.17 | < .001 |
| WMHv, normalized (log) | - | - | - | - | - | - | 1.25 | 1.20, 1.29 | < .001 |
| CMBs, superficial (1+) | - | - | - | - | - | - | 1.45 | 1.31, 1.61 | < .001 |
| CMBs, deep (1+) | - | - | - | - | - | - | 0.88 | 0.74, 1.04 | .120 |
| Infarction (1+) | - | - | - | - | - | - | 1.01 | 0.89, 1.15 | .900 |

\*OR = Odds Ratio, †CI = Confidence Interval. N = 1,352.
See **Supplementary Table 2** for description of nested model steps.
See **Supplementary Table 3** for nested models by cohort.

Repeating this stepwise analysis across cognitive stages, model parsimony improved when CVD biomarkers, but not MTLv, were added to the work-up of preclinical AD in the CU cohort (Δ AIC = –5.53, d_3_[4] = 3.22, p = .012). However, there was no consistent further benefit to prediction of Aβ-PET positivity for the later disease stages when either MTLv or CVD biomarkers were added (**Supplementary Tables 3and 4**). Regardless, for the combined accessible model (i.e. ‘Step 3’: Base + MTLv + CVD), we observed moderate to large effect sizes for the prediction of Aβ-PET positivity across all cohorts, with pseudo-R^2^ values of .24, .37, and .45 in the CU, MCI, and DEM cohorts, respectively (**Figure 2**).

**Figure 2:**
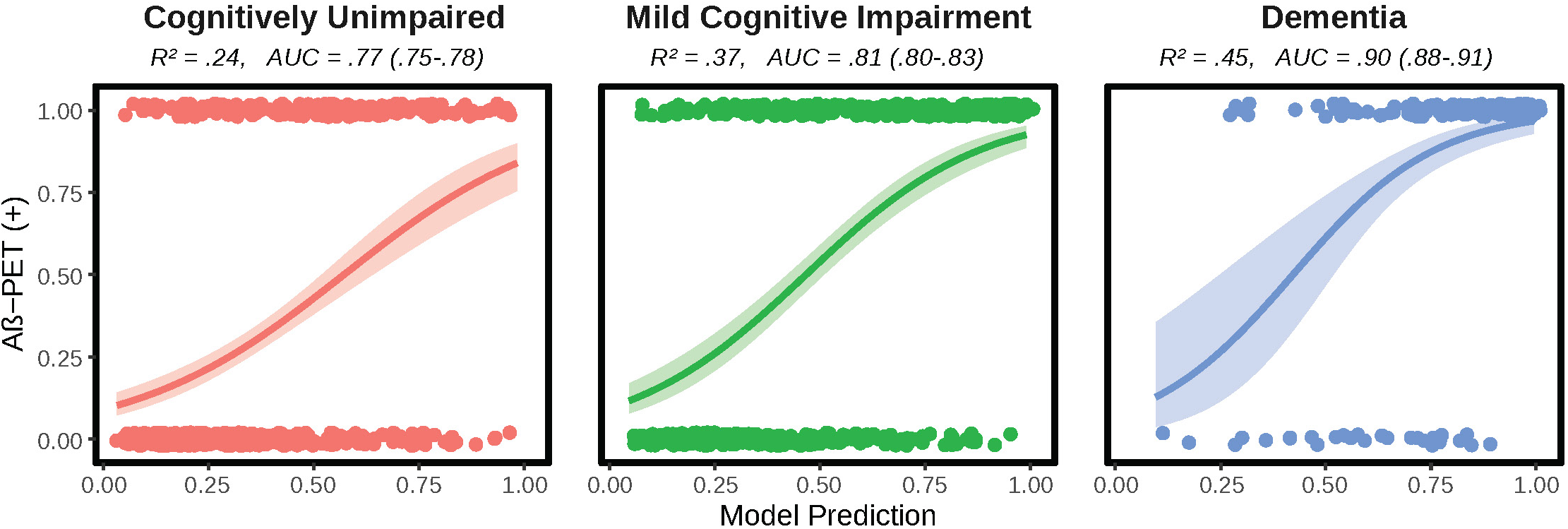
Combined logistic regression models by cohort. Logistic regression plots demonstrating statistically significant prediction of Aβ-PET status for each cohort, including cognitively unimpaired participants (N = 611), participants with mild cognitive impairment (N = 531) and participants with dementia (N = 210). Model predictions are plotted on the x-axis, while Aβ-PET status is plotted on the y-axis. Pseudo-R^2^ values and AUC values with a 95% CI are listed for each cohort.

### 3.7 Diagnostic performance of the combined accessible model by cohort

We finally sought to explore the diagnostic performance of our combined accessible model in discriminating between Aβ-positive and –negative individuals with ROC analysis, using Aβ-PET as the standard-of-truth. In the CU cohort, we identified a model AUC of .77 (95% CI: .75 – .78), consistent with moderate discriminatory ability between Aβ-PET positives and negatives using the predicted probabilities estimated by the model. In the MCI cohort, we identified slightly improved discrimination between Aβ-PET positives and negatives (AUC = .81 (95% CI: .80 – .83); in the DEM cohort, the model exhibited the highest discriminatory ability between Aβ-PET positives and negatives (AUC = .90 [95% CI: .88 – .91]). Additional data on the diagnostic performance of the combined accessible model, including ROC curves and histograms with fitted probability values are displayed in **Supplementary** Figure 1. Detailed analysis of the diagnostic performance for the ‘optimal’ probability cutoffs are listed for each cohort in **Supplementary Table 5.**

## 4. DISCUSSION

In recent years, there has been a growing emphasis on multimodal diagnostic approaches in AD, integrating traditional clinical measures with a range of biomarkers to enhance diagnostic accuracy. Our study extends this paradigm by incorporating accessible biomarkers of CVD into a model that already includes baseline demographics (age, sex), cognitive screening (MoCA), genetic information (APOE genotyping), along with clinically accessible neuroimaging measures of brain atrophy (MTL volume). Our study emphasizes the potential of comprehensive imaging assessments to provide a more holistic evaluation of patients, and highlights striking differences in the relationships between CVD biomarkers and Aβ neuropathology across the spectrum of cognitive decline.

Our findings highlight the need to consider each CVD biomarker distinctly in AD diagnostics and progression, a significant gap in the literature. For example, prior work indicates that the presence superficial “lobar” CMBs are highly specific to CAA [12]. Our finding of superficial CMBs’ uniformly positive association with Aβ-PET across cognitive stages aligns with prior studies and the Boston Neuroimaging Criteria 2.0, a widely accepted diagnostic strategy for CAA [12,30]. Deep CMBs (subcortical, periventricular, or brainstem) are less well characterized and may relate to mixed small vessel pathologies, including hypertensive vasculopathy [31,32]. However, we also found that deep CMBs displayed a biphasic relationship with Aβ-PET positivity, marked by a negative association in the CU cohort, and a positive one in later stages of MCI and dementia due to AD. While this requires further study, it is possible that mixed small vessel disease etiologies, affecting deeper brain regions, may explain the paradoxical positive association in MCI and dementia due to higher overall CVD burden contributing to clinically overt cognitive impairment [5,32–34].

Additionally, WMH has been increasingly recognized as a marker of small vessel disease and is posited to contribute to cognitive decline in AD. While mounting evidence links increased WMH volume to Aβ, tau and small vessel neuropathology, other studies suggest it may reflect AD-related Wallerian degeneration in later disease stages [21,35–37]. Our findings of a consistent association between WMH and Aβ-PET positivity across all cognitive stages suggest a stable and independent relationship with Aβ neuropathology. The reproducibility across cognitive stages, even after adjusting for other variables (including normalization of WMH to whole brain volume), further underlines WMH as a promising structural biomarker of Aβ pathology. This result echoes findings of longitudinal studies that link WMH to upstream Aβ accumulation and further progression of disease [38,39], rather than a downstream side-effect of Wallerian degeneration secondary to neurodegeneration associated with later disease stages. More work is needed to examine the performance of WMH alongside other CVD biomarkers in prediction of future disease progression.

Asymptomatic brain infarction is an understudied biomarker in AD and small vessel disease, due in part to challenges inherent in the variable size/distribution of pathology (cortical vs sub-cortical/lacunar), unclear timing of injury, as well as the underlying etiology (thrombotic, micro-embolic or watershed infarction) [13,19,40,41]. While we were not sufficiently powered to analyze these subtypes of stroke, or able to determine timing given the cross-sectional design and their ‘silent’ nature, taken together as a whole, we found that the presence of ischemic infarction was significantly associated with Aβ-PET positivity in preclinical AD, but not in MCI or dementia groups. Whereas longitudinal analysis in a stroke-specific cohort would be required to assess temporal relationships to Aβ positivity, our findings in preclinical AD are in line with basic science research linking stroke to Aβ plaque deposition and impairment in perivascular Aβ clearance pathways as a potential driver of AD, rather than an incidental finding [11].

Our approach aligns with the broader shift in AD research towards precision medicine, where individualized assessment strategies based on genetic, biomarker, and clinical data are crucial [2,42]. These results may facilitate the development of new CVD-based assessments to aid in detection of Aβ neuropathology during preclinical and symptomatic stages, potentially increasing diagnostic confidence, especially important given the gravity of a dementia diagnosis, in mixed cases where vascular and AD pathologies commonly co-exist [3–5]. Since MRI is already widely used in the clinical work-up of cognitive complaints [15], pending validation, these CVD biomarkers could be translated to inform work-up with blood biomarkers, Aβ-PET or cerebrospinal fluid analysis [43–45].

Prior to translation to clinical use, more work is needed to validate this model in a dataset that includes cases of vascular dementia alongside cases of “mixed” pathology and “pure” AD. This could enable development of AD-specific cutoffs for each CVD biomarker; for example, milder CVD biomarker changes may be more consistent with AD, while more severe increases in vascular burden could be more consistent with a primary diagnosis of vascular cognitive impairment. Based on our analysis, these optimal cutoffs may differ based on cognitive stage. While additional studies are needed to address differentiation from vascular dementia, our holistic approach aligns with the emerging trend in AD research, advocating for comprehensive diagnostic models that combine vascular markers into established research criteria such as the National Institute on Aging – Alzheimer’s Association Amyloid/Tau/Neurodegeneration Framework [2].

Our study has some important limitations, including potential ascertainment bias in the ADNI dataset due to a high prevalence of Aβ positivity, which does not fully represent the spectrum of vascular or other non-AD pathologies seen in clinical populations [4,40]. While excluding individuals with clear non-AD pathology likely increases our analysis’ specificity, it may limit its generalizability to clinical populations with mixed pathologies. Survival bias may also be at play, given that cerebral and systemic vascular diseases are a leading cause of morbidity and mortality in older populations, potentially contributing to underestimation of the impacts of ischemic stroke in later disease stages [46,47]. Furthermore, the ADNI dataset’s relative lack of racial or ethnic diversity limits our findings, as neuropathological studies show a higher vascular co-pathology burden in minoritized or traditionally underrepresented racial and ethnic groups [3]. Taken together, these limitations may lead to an underestimation of the relationship between these CVD biomarkers and Aβ positivity; more work is needed to longitudinally assess these relationships and better establish causality.

Future research should focus on several key areas to build upon our findings. Longitudinal studies are necessary to observe the progression of Aβ/tau pathology, neurodegeneration, and cognitive/functional decline over time in patients stratified by CVD biomarker status. As described above, more work is also needed to identify the accuracy of these biomarkers against vascular cognitive impairment and other types of dementia. These approaches could help define the bounds of AD, mixed vascular-AD dementia, and primary vascular cognitive impairment. By gaining deeper insights into the trajectory of AD, it may also lead to improved prediction of outcomes with novel amyloid-lowering immunotherapies, including risk of amyloid-related imaging abnormalities, a vascular consequence that underscores the need to better understand the relationship between CVD and AD with important implications for treatment decisions [48,49].

In summary, our study highlights the complex interplay between CVD and AD, pointing towards a more nuanced understanding of AD progression that incorporates vascular pathology. While more work is needed to define the bounds of AD, mixed dementia, and vascular cognitive impairment, our unique findings in a relatively large observational cohort may help to refine diagnostic criteria for AD and informs future studies in more racially diverse and diagnostically complex cohorts. If successful, this integrative approach could lead to more personalized and effective treatment strategies, improving outcomes for individuals with AD.

## Supporting information

Supplemental Files (All)

## Data Availability

All data produced are available online at adni.loni.usc.edu

## 6. ACKNOWLEDGEMENTS/CONFLICTS/FUNDING SOURCES/CONSENT STATEMENT

## 6.1 Acknowledgements

MDH designed the analysis, wrote the initial drafts of the manuscript, assisted with the statistical analysis, and created all tables and figures. MRC and MM reviewed the ADNI study procedures, assisted with writing the Methods and Supplementary Methods sections, and reviewed the manuscript. ZJK performed all data cleaning, analyzed data, and reviewed the manuscript. SE assisted with designing the analysis, assisted with the literature review and revised the manuscript. JLR, EDH and SPS reviewed all results, revised the initial drafts, and assisted with writing the final drafts of the manuscript. HO provided input on designing the analysis, reviewed the ADNI neuroimaging analysis pipeline, provided input on the statistical analysis and assisted with writing the final drafts of the manuscript.

The authors would like to thank the ADNI leadership, investigators and participants, the staff at LONI, and the faculty and staff of the Center for Neuroscience at University of California at Davis, the University of California at San Francisco, and the Mayo Clinic Aging and Dementia Imaging Research Laboratory for sharing their neuroimaging analysis to make our study possible.

## 6.4 Consent Statement

Written informed consent was obtained from all participants or authorized representatives prior to the start of study procedures, with local Institutional Review Boards and Research Ethics Boards providing oversight for the study. The authors certify that the study was performed in accordance with the ethical standards as laid down in the 1964 Declaration of Helsinki and its later amendments. For more information, see adni.loni.usc.edu.

## HIGHLIGHTS

- Cerebrovascular biomarkers linked to Aβ in AD.
- WMH, CMBs reliably predict Aβ-PET positivity.
- Relationships with Aβ-PET vary by cognitive stage.
- Novel accessible model predicts Aβ-PET status.
- Study supports multimodal diagnostic approaches.

## RESEARCH IN CONTEXT

- **Systematic Review:** Our comprehensive review of the Alzheimer disease (AD) radio-pathologic literature underscored that while cerebrovascular biomarkers are well-documented in small vessel disease, their individual and potentially additive effects in detecting amyloidosis are understudied. This highlighted the need for a focused study on how these biomarkers operate across different stages of AD, particularly as part of a multimodal diagnostic strategy.
- **Interpretation:** Our research newly demonstrates the unique and independent effects of cerebrovascular biomarkers in detecting amyloidosis in various cognitive stages of AD. This contribution improves the current understanding of specific cerebrovascular biomarkers across the AD spectrum, revealing nuanced relationships between these biomarkers and the disease.
- **Future Directions:** Future studies should seek to examine how location-specific cerebrovascular factors affect cognitive and functional decline over time, explore integration with blood biomarkers to enhance diagnostic accuracy, and establish their validity across diverse populations in cohorts with vascular and other non-AD dementias.

