## Supplemental Files (All) for "Utility of cerebrovascular imaging biomarkers to detect cerebral amyloidosis"

**Data used in preparation of this article were obtained from the Alzheimer’s Disease Neuroimaging Initiative (ADNI) database (adni.loni.usc.edu). As such, the investigators within the ADNI contributed to the design and implementation of ADNI and/or provided data but did not participate in analysis or writing of this report. A complete listing of ADNI investigators can be found at: <http://adni.loni.usc.edu/wp-content/uploads/how_to_apply/ADNI_Acknowledgement_List.pdf>

**Declaration of Competing Interests:** *Dr. Salloway has provided consultation to Biogen, Eisai, Avid, Lilly, Genentech, and Roche. Butler Hospital has received research grants from Biogen, Eisai, Avid, Roche, Genentech, Janssen and Lilly. The authors have no other competing interests to disclose.*

**SUPPLEMENTARY METHODS**

**Freesurfer measurements**

Cortical reconstruction and volumetric segmentation was performed with the Freesurfer image analysis suite, which is documented and freely available for download online (<http://surfer.nmr.mgh.harvard.edu/>). The technical details of these procedures are described in prior publications (Dale et al., 1999; Dale and Sereno, 1993; Fischl and Dale, 2000; Fischl et al., 2001; Fischl et al., 2002; Fischl et al., 2004a; Fischl et al., 1999a; Fischl et al., 1999b; Fischl et al., 2004b; Han et al., 2006; Jovicich et al., 2006; Segonne et al., 2004, Reuter et al. 2010, Reuter et al. 2012). Briefly, this processing includes motion correction and averaging (Reuter et al. 2010) of multiple volumetric T1 weighted images (when more than one is available), removal of non-brain tissue using a hybrid watershed/surface deformation procedure (Segonne et al., 2004), automated Talairach transformation, segmentation of the subcortical white matter and deep gray matter volumetric structures (including hippocampus, amygdala, caudate, putamen, ventricles) (Fischl et al., 2002; Fischl et al., 2004a) intensity normalization (Sled et al., 1998), tessellation of the gray matter white matter boundary, automated topology correction (Fischl et al., 2001; Segonne et al., 2007), and surface deformation following intensity gradients to optimally place the gray/white and gray/cerebrospinal fluid borders at the location where the greatest shift in intensity defines the transition to the other tissue class (Dale et al., 1999; Dale and Sereno, 1993; Fischl and Dale, 2000). Once the cortical models are complete, a number of deformable procedures can be performed for further data processing and analysis including surface inflation (Fischl et al., 1999a), registration to a spherical atlas which is based on individual cortical folding patterns to match cortical geometry across subjects (Fischl et al., 1999b), parcellation of the cerebral cortex into units with respect to gyral and sulcal structure (Desikan et al., 2006; Fischl et al., 2004b), and creation of a variety of surface based data including maps of curvature and sulcal depth. This method uses both intensity and continuity information from the entire three dimensional MR volume in segmentation and deformation procedures to produce representations of cortical thickness, calculated as the closest distance from the gray/white boundary to the gray/CSF boundary at each vertex on the tessellated surface (Fischl and Dale, 2000). The maps are created using spatial intensity gradients across tissue classes and are therefore not simply reliant on absolute signal intensity. The maps produced are not restricted to the voxel resolution of the original data thus are capable of detecting submillimeter differences between groups. Procedures for the measurement of cortical thickness have been validated against histological analysis (Rosas et al., 2002) and manual measurements (Kuperberg et al., 2003; Salat et al., 2004). Freesurfer morphometric procedures have been demonstrated to show good test-retest reliability across scanner manufacturers and across field strengths (Han et al., 2006; Reuter et al., 2012).

**Aseg Atlas Information**

The aseg atlas is built from 40 subjects acquired using the same mp-rage sequence (by people at Wash U ages ago in collaboration with Randy Buckner). The subjects that make up the atlas are distributed in 4 groups of 10 subjects each: (1) young, (2) middle aged, (3) healthy older adults, (4) older adults with AD.

**Image acquisition**

T1, T2, FLAIR and T2* weighted (GRE) imaging sequences, without contrast, were performed across 3-tesla field strength Siemens, Philips, and GE scanners using standardized protocols as previously described (Jack et al., 2010; Weiner et al., 2017).

**Quantification of medial temporal lobe volume**

Volumetric T1 weighted MRI images were processed using the FreeSurfer Image Analysis Suite. Freesurfer was used for cortical reconstruction and volumetric segmentation using the 2010 Desikan-Killany atlas (Desikan et al., 2006). Before being modified by the FreeSurfer program, T1 weighted images, in NiFTI format were pre-processed by the Mayo Clinic. MTL volume was determined using a high-resolution coronal T2 MRI sequence. Cortical reconstruction and volume reconstruction were performed with Freesurfer.

**Quantification of white matter hyperintensities**

White matter hyperintensity (WMH) volumes were determined by using a segmentation of the high-resolution 3D T1 and FLAIR sequences. Using an automated atlas, non-brain structures were removed from the 3DT1 images, the corresponding FLAIR image transformed to the image and the 3D T1 image aligned as previously described (Decarli et al., 1996; Fletcher et al., 2012). Estimation of WMH was performed using a modified probability structure, with probability likelihoods of WMH in each voxel generated and a binary WMH mask back transformed to calculate tissue volume (DeCarli et al., 1999).

**Quantification of cerebral microbleeds**

Cerebral microbleeds (CMBs) were defined as focal hypodensities visible in T2* GRE images. Location was determined based on corresponding T1-weighted images. CMBs were confirmed as “definite” or “probable” based upon context across multiple images, then classified based on location as superficial (lobar cortical and cerebellar regions) and deep (subcortical, periventricular white matter and brainstem regions) as previously described (Charidimou et al., 2022).

**Quantification of infarcts**

Infarcts were determined by a physician specifically trained in interpretation of MRI. Infarct size, location and other imaging characteristics were recorded and confirmed by cross-checking against CSF density on T1 sequences and distinct separation from vessels in certain areas of the brain as previously described (DeCarli et al., 2005). Only lesions equal to or greater than 3mm in size were considered cerebral infarcts for this analysis.

**Positron emission tomography**

18F-Florbetaben and 18F-Florbetapir Aβ-PET imaging was conducted under the respective ADNI protocols. Scan parameters for 18F-Florbetaben consisted of: 300 MBq (8.1 mCi); four 5-minute frames acquired 90 minutes post-injection. Scan parameters for 18F-Florbetapir consisted of: 370 MBq (10.0 mCi); four 5-minute frames acquired 50 minutes post-injection) (Royse et al., 2021). PET scans were co-registered to the MRI closest in time, and mean tracer uptake within an MRI-defined cortical summary region (frontal, anterior/posterior cingulate, lateral parietal, lateral temporal) and reference region (whole cerebellum). Tracer intensity in the cortical summary region was divided by the reference region and expressed as a Standard Uptake Value Ratio (SUVR). Aβ-PET positivity was defined as SUVR> 1.11 and SUVR > 1.08 for florbetapir and florebetapen, respectively (Landau et al., 2014, 2013). For quantitative comparisons, SUVR was transformed to a standardized centiloid scale as previously described (Royse et al., 2021).

| **Supplementary Table 1: Extended demographics and clinical characteristics** | | | |
| --- | --- | --- | --- |
| **Characteristic** | **Overall**, N = 1,352*^1^* | **Aβ-**, N = 679*^1^* | **Aβ+**, N = 673*^1^* |
| Age, years | 72 (7) | 71 (7) | 73 (7) |
| *Unknown* | *3* | *0* | *3* |
| Sex, male | 674 (50%) | 342 (50%) | 332 (49%) |
| Education, years | 16.39 (2.50) | 16.62 (2.38) | 16.15 (2.60) |
| APOE4, alleles |  |  |  |
| *0* | *751 (56%)* | *516 (77%)* | *235 (35%)* |
| *1* | *467 (35%)* | *139 (21%)* | *328 (49%)* |
| *2* | *116 (8.7%)* | *11 (1.7%)* | *105 (16%)* |
| *Unknown* | *18* | *13* | *5* |
| MoCA, score | 23.4 (4.4) | 24.9 (3.3) | 22.0 (4.8) |
| *Unknown* | *25* | *10* | *15* |
| MTLv, normalized (log) | -4.25 (0.12) | -4.22 (0.10) | -4.29 (0.13) |
| *Unknown* | *176* | *88* | *88* |
| WMHv, normalized (log) | -6.34 (1.39) | -6.68 (1.36) | -6.00 (1.34) |
| *Unknown* | *12* | *7* | *5* |
| CMBs, present |  |  |  |
| *Any location (1+)* | *363 (27%)* | *148 (22%)* | *215 (32%)* |
| *Superficial (1+)* | *301 (22%)* | *111 (17%)* | *190 (28%)* |
| *Deep (1+)* | *95 (7.1%)* | *40 (6.0%)* | *55 (8.2%)* |
| *Unknown* | *11* | *7* | *4* |
| Infarct, present (1+) | 94 (8.1%) | 42 (7.3%) | 52 (8.8%) |
| *Unknown* | *192* | *107* | *85* |
| Clinical diagnosis |  |  |  |
| *Cognitively normal* | *611 (45%)* | *413 (61%)* | *198 (29%)* |
| *Mild cognitive impairment* | *531 (39%)* | *237 (35%)* | *294 (44%)* |
| *Dementia* | *210 (16%)* | *29 (4.3%)* | *181 (27%)* |
| Aβ-PET burden, centiloids | 37 (42) | 3 (10) | 72 (34) |
| *^1^* n (%); Mean (SD); Median (IQR) | | | |

| **Supplementary Table 2: Nested model components** | | |
| --- | --- | --- |
| **Model 1 (Base)** | **Model 2 (Base + *MTLv^1^*)** | **Model 3 (Base + MTLv^1^ + *CVD^2^*)** |
| Age | Age | Age |
| Sex | Sex | Sex |
| APOE4 | APOE4 | APOE4 |
| MoCA | MoCA | MoCA |
| - | *MTLv* | MTLv |
| - | - | *WMHv* |
| - | - | *Superficial CMBs* |
| - | - | *Deep CMBs* |
| - | - | *Ischemic infarction* |
| ^1^MTLv refers to the mean volume of the medial temporal lobes, normalized to whole brain volume and expressed as a log-transformed value. ^2^Cerebrovascular disease (CVD) variables includes white matter hyperintensity volume (WMHv), cerebral microbleeds (CMBs) and ischemic infarction. WMHv was normalized to whole brain volume and expressed as a log-transformed value. Based on atlas location, CMBs were divided into superficial (occurring in cortical and cerebellar areas) and deep (occurring in subcortical, periventricular and brainstem areas). | | |

| **Supplementary Table 3: Adjusted odds ratios for nested models** | | | | | | | | | |
| --- | --- | --- | --- | --- | --- | --- | --- | --- | --- |
|  | **Model 1 (Base)** | | | **Model 2 (Base + MTL)** | | | **Model 3 (Base + MTL + CVD)** | | |
| **Characteristic** | **OR***^1^* | **95% CI^2^** | **p-value** | **OR***^1^* | **95% CI***^2^* | **p-value** | **OR***^1^* | **95% CI***^2^* | **p-value** |
| Overall Cohort | | | | | | | | | |
| Age, years | 1.08 | 1.07, 1.08 | <0.001 | 1.07 | 1.06, 1.08 | <0.001 | 1.05 | 1.04, 1.06 | <0.001 |
| Sex, male | 0.67 | 0.62, 0.73 | <0.001 | 0.66 | 0.60, 0.71 | <0.001 | 0.66 | 0.60, 0.71 | <0.001 |
| APOE4, alleles | 5.70 | 5.29, 6.15 | <0.001 | 5.63 | 5.22, 6.08 | <0.001 | 5.66 | 5.24, 6.11 | <0.001 |
| MoCA, score | 0.86 | 0.85, 0.87 | <0.001 | 0.89 | 0.88, 0.90 | <0.001 | 0.89 | 0.88, 0.90 | <0.001 |
| MTLv, normalized (log) | - | - | - | 0.10 | 0.07, 0.15 | <0.001 | 0.12 | 0.08, 0.17 | <0.001 |
| WMHv, normalized (log) | - | - | - | - | - | - | 1.25 | 1.20, 1.29 | <0.001 |
| CMBs, superficial (1+) | - | - | - | - | - | - | 1.45 | 1.31, 1.61 | <0.001 |
| CMBs, deep (1+) | - | - | - | - | - | - | 0.88 | 0.74, 1.04 | 0.120 |
| Infarction (1+) | - | - | - | - | - | - | 1.01 | 0.89, 1.15 | 0.900 |
| CU Cohort | | | | | | | | | |
| Age, years | 1.11 | 1.10, 1.12 | <0.001 | 1.11 | 1.10, 1.12 | <0.001 | 1.09 | 1.08, 1.10 | <0.001 |
| Sex, male | 0.58 | 0.51, 0.65 | <0.001 | 0.57 | 0.51, 0.65 | <0.001 | 0.57 | 0.50, 0.65 | <0.001 |
| APOE4, alleles | 3.91 | 3.49, 4.39 | <0.001 | 3.91 | 3.49, 4.39 | <0.001 | 3.98 | 3.55, 4.48 | <0.001 |
| MoCA, score | 0.98 | 0.96, 1.00 | 0.076 | 0.98 | 0.96, 1.00 | 0.086 | 0.98 | 0.95, 1.00 | 0.074 |
| MTLv, normalized (log) | - | - | - | 0.80 | 0.44, 1.47 | 0.500 | 1.07 | 0.58, 1.99 | 0.800 |
| WMHv, normalized (log) | - | - | - | - | - | - | 1.25 | 1.19, 1.31 | <0.001 |
| CMBs, superficial (1+) | - | - | - | - | - | - | 1.38 | 1.18, 1.62 | <0.001 |
| CMBs, deep (1+) | - | - | - | - | - | - | 0.51 | 0.39, 0.66 | <0.001 |
| Infarction (1+) | - | - | - | - | - | - | 1.31 | 1.09, 1.58 | 0.005 |
| MCI Cohort | | | | | | | | | |
| Age, years | 1.07 | 1.06, 1.08 | <0.001 | 1.06 | 1.05, 1.07 | <0.001 | 1.05 | 1.04, 1.06 | <0.001 |
| Sex, male | 0.79 | 0.69, 0.90 | <0.001 | 0.77 | 0.68, 0.88 | <0.001 | 0.79 | 0.70, 0.91 | <0.001 |
| APOE4, alleles | 6.58 | 5.86, 7.41 | <0.001 | 6.55 | 5.83, 7.38 | <0.001 | 6.62 | 5.88, 7.48 | <0.001 |
| MoCA, score | 0.93 | 0.91, 0.95 | <0.001 | 0.94 | 0.92, 0.96 | <0.001 | 0.95 | 0.93, 0.97 | <0.001 |
| MTLv, normalized (log) | - | - | - | 0.09 | 0.05, 0.16 | <0.001 | 0.09 | 0.05, 0.16 | <0.001 |
| WMHv, normalized (log) | - | - | - | - | - | - | 1.15 | 1.09, 1.21 | <0.001 |
| CMBs, superficial (1+) | - | - | - | - | - | - | 1.37 | 1.17, 1.61 | <0.001 |
| CMBs, deep (1+) | - | - | - | - | - | - | 1.32 | 1.02, 1.72 | 0.037 |
| Infarction (1+) | - | - | - | - | - | - | 0.89 | 0.73, 1.08 | 0.200 |
| Dementia Cohort | | | | | | | | | |
| Age, years | 1.02 | 1.00, 1.03 | 0.066 | 1.01 | 0.99, 1.03 | 0.200 | 0.99 | 0.97, 1.01 | 0.200 |
| Sex, male | 0.26 | 0.18, 0.36 | <0.001 | 0.23 | 0.16, 0.33 | <0.001 | 0.20 | 0.14, 0.29 | <0.001 |
| APOE4, alleles | 9.97 | 7.45, 13.6 | <0.001 | 10.30 | 7.65, 14.1 | <0.001 | 10.50 | 7.73, 14.7 | <0.001 |
| MoCA, score | 0.89 | 0.86, 0.92 | <0.001 | 0.92 | 0.89, 0.96 | <0.001 | 0.93 | 0.89, 0.96 | <0.001 |
| MTLv, normalized (log) | - | - | - | 0.01 | 0.00, 0.03 | <0.001 | 0.01 | 0.00, 0.03 | <0.001 |
| WMHv, normalized (log) | - | - | - | - | - | - | 1.54 | 1.33, 1.79 | <0.001 |
| CMBs, superficial (1+) | - | - | - | - | - | - | 2.17 | 1.46, 3.29 | <0.001 |
| CMBs, deep (1+) | - | - | - | - | - | - | 2.99 | 1.36, 7.33 | 0.011 |
| Infarction (1+) | - | - | - | - | - | - | 1.28 | 0.74, 2.28 | 0.400 |
| *^1^* OR = Odds Ratio, ^2^CI = Confidence Interval | | | | | | | | | |

| **Supplementary Table 4: Change in nested model goodness-of-fit with additional predictors** | | | | | | |
| --- | --- | --- | --- | --- | --- | --- |
| **Model^1^** | **Nagelkerke R^2^** | **AIC^2^** | **Δ AIC^3^** | **Λ^4^** | **d_3_ (df)^5^** | **p-value^6^** |
| Overall Cohort | | | | | | |
| Step 1 (Base) | .38 | 1,425.22 | - | - | - | - |
| Step 2 (Base + MTLv) | .39 | 1,413.64 | -11.43 | 13.43 | 11.33 (1) | .001 |
| Step 3 (Base + MTLv + CVD) | .41 | 1,397.46 | -15.78 | 23.78 | 5.30 (4) | < .001 |
| CU Cohort | | | | | | |
| Step 1 (Base) | .21 | 677.64 | - | - | - | - |
| Step 2 (Base + MTLv) | .22 | 679.52 | 1.95 | 0.05 | 0.05 (1) | .828 |
| Step 3 (Base + MTLv + CVD) | .24 | 673.82 | -5.53 | 13.53 | 3.22 (4) | .012 |
| MCI Cohort | | | | | | |
| Step 1 (Base) | .35 | 582.71 | - | - | - | - |
| Step 2 (Base + MTLv) | .36 | 574.40 | -4.11 | 6.11 | 5.53 (1) | .019 |
| Step 3 (Base + MTLv + CVD) | .37 | 576.41 | +2.28 | 5.71 | 1.32 (4) | .261 |
| Dementia Cohort | | | | | | |
| Step 1 (Base) | .36 | 132.24 | - | - | - | - |
| Step 2 (Base + MTLv) | .40 | 128.87 | -2.86 | 4.86 | 3.00 (1) | .094 |
| Step 3 (Base + MTLv + CVD) | .45 | 129.14 | 0.60 | 7.40 | 1.68 (4) | .153 |
| ^1^Nested regression model for each cohort was developed with forced forward selection of predefined predictor variables; ^2^Akaike Information Criterion (AIC) for nested model (absolute); ^3^Change in AIC value from previous step; ^4^Likelihood Ratio Test (LRT) statistic (λ) for constrained model compared to previous step; ^5^d_3_ statistic indicating goodness-of-fit for imputed data, with degrees of freedom (df) for number of added predictors; ^6^Statistical significance of improvement in nested model goodness-of-fit (d_3_) compared to previous step, adjusted for between-imputation variance. | | | | | | |

| **Supplementary Table 5: Diagnostic performance of nested models at Youden’s cut-off** | | | | |
| --- | --- | --- | --- | --- |
| **Model** | **Probability Cutoff** | **Accuracy** | **Sensitivity** | **Specificity** |
| Overall Cohort | | | | |
| Step 1 (Base) | 0.48 | 73.95% | 72.87% | 75.02% |
| Step 2 (Base + MTLv) | 0.47 | 74.36% | 74.23% | 74.49% |
| Step 3 (Base + MTLv + CVD) | 0.51 | 75.34% | 72.01% | 78.65% |
| CU Cohort | | | | |
| Step 1 (Base) | 0.27 | 67.1% | 77.78% | 61.99% |
| Step 2 (Base + MTLv) | 0.28 | 67.09% | 76.72% | 62.47% |
| Step 3 (Base + MTLv + CVD) | 0.32 | 71.9% | 71.87% | 71.91% |
| MCI Cohort | | | | |
| Step 1 (Base) | 0.61 | 73.52% | 66.12% | 82.7% |
| Step 2 (Base + MTLv) | 0.66 | 73.22% | 62.89% | 86.03% |
| Step 3 (Base + MTLv + CVD) | 0.60 | 74.76% | 68.03% | 83.12% |
| Dementia Cohort | | | | |
| Step 1 (Base) | 0.88 | 76.52% | 73.87% | 93.1% |
| Step 2 (Base + MTLv) | 0.90 | 73.76% | 70.88% | 91.72% |
| Step 3 (Base + MTLv + CVD) | 0.90 | 76.95% | 74.81% | 90.34% |
| ROC analysis of logistic regression fitted values were used to calculate Youden’s optimal cut-off, deriving the predicted probability of Aβ-PET positivity. Corresponding diagnostic accuracy, sensitivity and specificity is listed. | | | | |

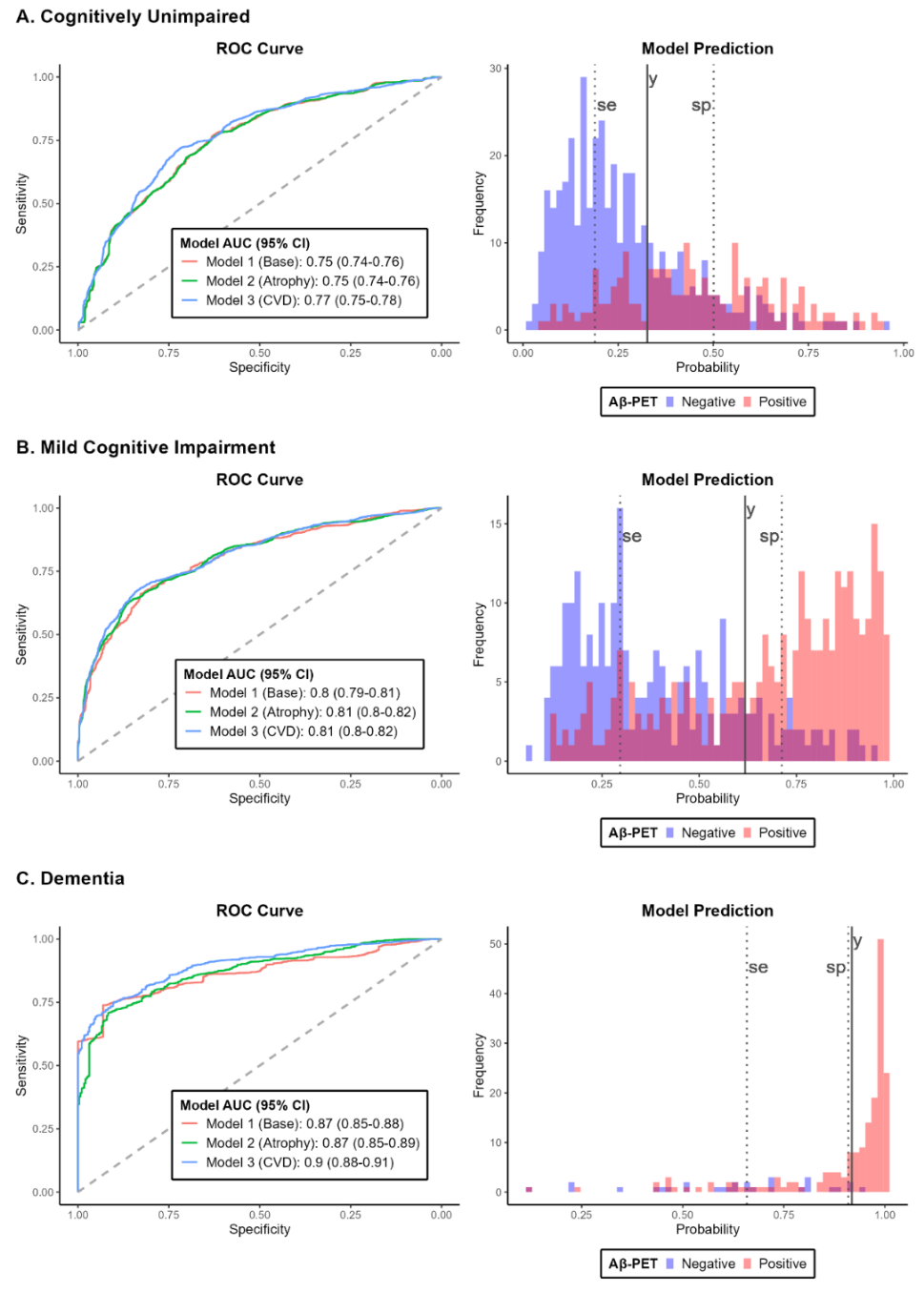

**Supplementary Figure 1: Diagnostic performance of nested and combined models by cohort.** ROC curves (left) show prediction of Aβ-PET status across cognitive stages. Addition of CVD-related MRI measures (blue line) improved diagnostic performance in CU participants, while addition of atrophy measures (MTLv) improved performance in MCI participants. Histograms (right) show distribution of combined model predictions, colored by Aβ-PET status (negative – blue, positive – red), and overlayed with lines denoting specific cut-offs identified by our analysis. Solid lines (y) represent the Youden index and dashed lines represent cut-offs with 90% sensitivity (se) and specificity (sp). CU: Cognitively unimpaired (N = 611), MCI: Mild cognitive impairment (N = 531) or DEM: Dementia (N = 210).

**ACKNOWLEDGEMENTS, CONFLICTS AND FUNDING SOURCES**

**Acknowledgements:** MDH designed the analysis, wrote the initial drafts of the manuscript, assisted with the statistical analysis, and created all tables and figures. MRC and MM reviewed the ADNI study procedures, assisted with writing the Methods and Supplementary Methods sections, and reviewed the manuscript. ZJK performed all data cleaning, analyzed data, and reviewed the manuscript. SE assisted with designing the analysis, assisted with the literature review and revised the manuscript. JLR, EDH and SPS reviewed all results, revised the initial drafts, and assisted with writing the final drafts of the manuscript. HO provided input on designing the analysis, reviewed the ADNI neuroimaging analysis pipeline, provided input on the statistical analysis and assisted with writing the final drafts of the manuscript.

The authors would like to thank the ADNI leadership, investigators and participants, the staff at LONI, and the faculty and staff of the Center for Neuroscience at University of California at Davis, the University of California at San Francisco, and the Mayo Clinic Aging and Dementia Imaging Research Laboratory for sharing their neuroimaging analysis to make our study possible.

**Competing Interests:** M. Howe: *None to disclose.* M. Caruso: *None to disclose.* M. Manoochehri: *None to disclose.* Z. Kunicki: *None to disclose.* S. Emrani: *None to disclose.* J. Rudolph: *None to disclose.* E. Huey: *None to disclose.* S. Salloway: *Dr. Salloway has provided consultation to Biogen, Eisai, Avid, Lilly, Genentech, and Roche.* H. Oh: *None to disclose. Butler Hospital has received research grants from Biogen, Eisai, Avid, Roche, Genentech, Janssen and Lilly.*

**Funding:** Salary support to M. Howe is provided by NIMH 2R25MH101076-06A1 (Audrey Tyrka, PI). S. Salloway is supported by NIA 2P01AG051449-06 (John Sedivy, PI). H. Oh is supported by NIA R01AG068990 (Hwamee Oh, PI). Data collection and sharing for this project was funded by the Alzheimer's Disease Neuroimaging Initiative (ADNI) (National Institutes of Health Grant U01 AG024904) and DOD ADNI (Department of Defense award number W81XWH-12-2-0012). ADNI is funded by the National Institute on Aging, the National Institute of Biomedical Imaging and Bioengineering, and through generous contributions from the following: AbbVie, Alzheimer’s Association; Alzheimer’s Drug Discovery Foundation; Araclon Biotech; BioClinica, Inc.; Biogen; Bristol-Myers Squibb Company; CereSpir, Inc.; Cogstate; Eisai Inc.; Elan Pharmaceuticals, Inc.; Eli Lilly and Company; EuroImmun; F. Hoffmann-La Roche Ltd and its affiliated company Genentech, Inc.; Fujirebio; GE Healthcare; IXICO Ltd.; Janssen Alzheimer Immunotherapy Research & Development, LLC.; Johnson & Johnson Pharmaceutical Research & Development LLC.; Lumosity; Lundbeck; Merck & Co., Inc.; Meso Scale Diagnostics, LLC.; NeuroRx Research; Neurotrack Technologies; Novartis Pharmaceuticals Corporation; Pfizer Inc.; Piramal Imaging; Servier; Takeda Pharmaceutical Company; and Transition Therapeutics. The Canadian Institutes of Health Research is providing funds to support ADNI clinical sites in Canada. Private sector contributions are facilitated by the Foundation for the National Institutes of Health (www.fnih.org). The grantee organization is the Northern California Institute for Research and Education, and the study is coordinated by the Alzheimer’s Therapeutic Research Institute at the University of Southern California. ADNI data are disseminated by the Laboratory for Neuro-Imaging at the University of Southern California.
